# Medical Errors Among Postgraduate Medical Students Joining The Iraqi and Arab Councils For Medical Specializations - Medical City Complex

**DOI:** 10.1101/2023.12.24.23300506

**Authors:** Deema Thamer Mahmood, Niam Kamal Kareem, Mays Mohammed Abed, Ali M.J. Al-mothaffar

## Abstract

**Background:** All humans make mistakes. Unintentional medical mistakes affect patients and their families, but they can also have a negative behavioral, psychological and emotional impact on the involved healthcare practitioner or providers. These could include emotional distress, fear, feeling guilt and even depression.

**Aim:** This study aims to describe the events contributing to medical errors and the impact of these errors on behavior among postgraduate medical students joining the Iraqi and Arabic councils for medical specialty.

**Subjects and Methods:** A cross-sectional study was conducted in Medical city complex, Baghdad, Iraq, by convenient sampling. Data were collected by a self-administered questionnaire form, which was given to all participants and retrieved at the same day. The questionnaire contains three sections (13 items), in the first section it mainly asks about personal information of the participant and their specialty, Second section contains the main question of the study which is whether they have been involved in a medical error or not, and the third section asks about the events and behavior that are related to those errors like the time of the error, the most important contributing factor, the strongest feeling that they felt after this error and the most obvious behavioral response after the error.

**Results:** The total number of participants was 200 individuals. 111 of them were juniors and 89 were senior doctors. Gender (p-value< 0.0001) was significantly associated with medical errors, notably males tended to be involved in medical errors more than females with a number of 46 (40.7%) compared to 21 (24.13%) respectively. there is statistically significant association between decreased navigation of social media applications during work hours for non-work and medical error (P value < 0.0001). Furthermore, specialty was significantly associated with medical errors (P value < 0.0001) as surgical specialty tended to be involve in medical errors more than medical specialty with a number of 34 (38.2%) compared to 33(29.7%) respectively.

**Conclusion:** Distraction of attention was the most contributing factor behind medical errors. Emotional distress was the most common feeling experienced by participants. Increasing vigilance was the most behavioral response among participants following their involvement in medical errors. Furthermore, male doctors tend to be involved in medical errors more than female doctors. Surgical specialty tends to be involved in medical errors more than medical specialty.

## INTRODUCTION

Medical errors are unavoidable and can have catastrophic consequences for the patient, treating physician, nurses, and the facility. ^(1,2)^ Creating a safe healthcare system requires planning care procedures to protect patients from unintentional harm. It has been reported that medical errors cause up to 98,000 deaths annually in the United States, making them a greater cause of death than car crashes.^(3)^

Almost all physicians have made mistakes, but they frequently choose not to disclose them to patients or their families. Doctors’ error is a common occurrence in clinical practice, but it is underreported.^[4]^

Very little is known about the causes and effects of medical errors as a result of this underreporting. Furthermore, dealing with a medical error is never simple, which is why it is never disclosed.^(5)^ Even though it can be challenging to admit when you’ve made a mistake, you must confront it and make an effort to grow from it in order to avoid making the same mistakes again. Determining the risk factors for medical errors is an essential first step in preventing them and a key objective of ensuring the quality of care.^(6)^

One of the fundamental issues facing health systems worldwide is medication errors, which poses a major risk to patient safety ^(7,8)^. Errors with medication administration can have unpleasant outcomes, including extended hospital stays, higher treatment expenses, and even death ^(9)^. According to the findings of a survey done in the UK in 2018, medication errors cause complications for over 2 million people annually and result in approximately 100,000 deaths ^(10)^. In America, medication errors rank as the third most common cause of death ^(11)^. Medication errors can have a significant financial impact on the healthcare industry in addition to eroding patients’ confidence in doctors and the healthcare system ^(12)^

In Europe, 8–12% of hospital admissions are the result of medical errors, including adverse events connected to healthcare ^(10)^. At least half of the injuries suffered by hospitalized patients could be avoided ^(12)^.

Furthermore, it was noted that medication errors account for 100,000 hospital admissions per year and that at least one medication error occurs every day in United States ^(13,14)^. Medication errors (MEs) also affect 7% of inpatients on a daily basis; most of these patients are either in extended hospitalization care or are referred to critical care or high-dependency care units ^[15,16]^. Medication administration in such high-complexity situations like intensive care frequently requires extraordinary effort and interventions from healthcare providers ^(16)^

In both developed and developing nations, medical errors increase the cost of hospitalization and medical expenses, which lowers the quality of healthcare systems ^(17,18])^

Previous studies showed an increase in medical errors events despite the development of diagnostic and treatment services because of the complex issues of treatment and its timing ^(19-22)^. The prevalence of medical errors varied from 1 to 40%, per various studies. An estimated 17% of admissions to diagnostic and treatment facilities result in unfavorable outcomes ^(20-25)^

A 2013 review study done in 10 of 15 Middle Eastern countries including Iran and excluding Iraq found that error rates vary from 7% to 90% for medication prescription errors and 9% to 80 % for medication administration errors. ^(26)^

It is challenging to find a consistent cause of errors, and even if found, it is challenging to provide a consistent viable solution that minimizes the chances of a recurrent event. By recognizing untoward events that occur, learning from them, and working toward preventing them, patient safety can be improved ^(27)^

Medical errors might take place during diagnosis, giving prescriptions, surgery, employing medical equipment, preparing lab reports, and so on ^(28)^. However, medical errors in the American health system are due to improper communications, physicians’ errors in prescribing drugs, and problems in health information systems. In addition, factors such as large populations and inefficient and poor communication among clinical wards are effective in increasing the rate of medical errors ^(29)^. Under-staffed nurses and medical team members, workload, fatigue, and work pressure on medical team members were the most noted causes of medical errors by about half of the physicians ^(30)^. Poor education provided to health staff, stressful work conditions and increase of patient-to-nurse ratio were some of the reported causes of medicine-related errors ^(31)^.

This study aims to describe the events contributing to medical errors and the impact of these errors on behavior among postgraduate medical students joining the Iraqi and Arabic councils for medical specialty.

## SUBJECTS AND METHODS

It is a cross-sectional study was conducted at Medical City Complex during the period from June 2023 to December 2023.

Postgraduate Medical students joining the Iraqi and Arabic councils for Medical Specialization at Medical City Complex participated in this study.

A convenient sample of Iraqi and Arabic board candidates who are available at the time of data collection and agreed to participate.

The questionnaire form was prepared after reviewing the literatures and was modified by the researchers and supervisor then approved by five experts in the field.

- The questionnaire form contains 14 questions divided into three sections, section one includes questions about general information of participants such as age, gender, marital status, recent board program grade, year since graduation from medical college, average night sleeping hours, navigating social media during work hours and their specialty, meanwhile, section two includes a question about their involvement in medical errors. Section three contains questions about the participants’ ‘perception of cause of medical error, their responses to these errors, its disclosure and the effect of that error (constructive or defensive) on their behavior. if the participant was involved and about the disclosure of medical errors. These questions were presented to the participants as multiple-choice questions (11 questions) and close-ended questions (3 questions).
- To asses participant’s perception about the cause of error, we classified the causes into (Medication prescription error, procedural error, distraction of attention, knowledge gap, team communication error, lack of necessary equipment, using drug of sedation) after reviewing the literatures and modify it according to the study circumstances and researcher’s and supervisor viewpoints.
- To asses participant’s feelings in reaction to the error, we used a classification of (emotional distress, sadness, feeling guilty, inadequacy, frustration, fear, it was not my fault) depend on previous study in such topic^(32)^
- To assess participants’ behavioral response to errors, we used a classification of (information seeking, vigilance and defensive practice) depend on previous study in such topic.(32)
- The Self-administered questionnaire form was hand-distributed to all participants and retrieved on the same day.
- A Pilot study was conducted in March 2023 at AL-Yarmouk Teaching Hospital on 12 participants. The Response rate was 100%. Minor modification has been made to question 4 in the questionnaire form by adding a (from medical college) sentence after facing a misunderstanding from two participants. Another minor modification has been made to question 10 by adding (I can’t recall) choice because three participants couldn’t remember it. Last minor modification has been made to question 11 in the questionnaire form by making the sentence (choose one) in bold form because two participants have chosen more than one choice.

### Data Processing and Analysis

- Data were entered and processed using IBM SPSS 25 Statistics.
- Continuous variables were presented as mean ± standard deviation. Descriptive statistics in the form of frequency and relative frequency distribution tables were laid down. Statistical tests were used accordingly including Chi-square and student-t tests.
- Findings with a p-value<0.05 are considered statistically significant. For statistical purposes, the participants from the specialties of Internal Medicine, Clinical Hematology, Family Medicine, Rheumatology, Emergency, and Pediatrics have been combined under the term “Medical Specialty”.
- For statistical purposes, the participants from the specialties of General Surgery, Gynae-obstetrics, Orthopedics, Anesthesia, and Radiology have been combined under the term “Surgical Specialty”.
- For statistical purposes, participants from the first and second years of board program were combined under the term “Junior”.
- For statistical purposes, participants from the third, fourth, and fifth years of board program were combined under the term “Senior”.
- For statistical purposes, participants who are married, divorced, and widows were combined under the term “Married”.
- A number of 14 samples have been neglected due to invalid data and considered as exclusion criteria.
- A question about disclosing of medical errors has been removed due to its inappropriate placement in the questionnaire form caused many participants to dismiss it.
- Ethical approval was obtained from the Scientific Committee/College of Medicine/University of Baghdad.
- Before asking for their verbal consent, all subjects were given a brief description of the study’s objective, procedures, and value.
- The distributing and collecting process of the questionnaires was done by investigators who are unrelated to these objects in any manner, and all information was kept safe.
- During the data analysis process, identification numbers were used instead of names and personal data, which was protected and only accessible to the investigators.

## RESULTS

This study included 200 participants of post-graduate students who joined the Iraqi and Arabic councils for medical specializations at Medical City Complex.

Table (1) shows that most of the participants were at the age of 31.46+/-2.4 (26-44).Males accounted for 56.5% of participants, 61% were married and 55.55% were juniors and the same percentage were in medical specialty.

**Table 1:**
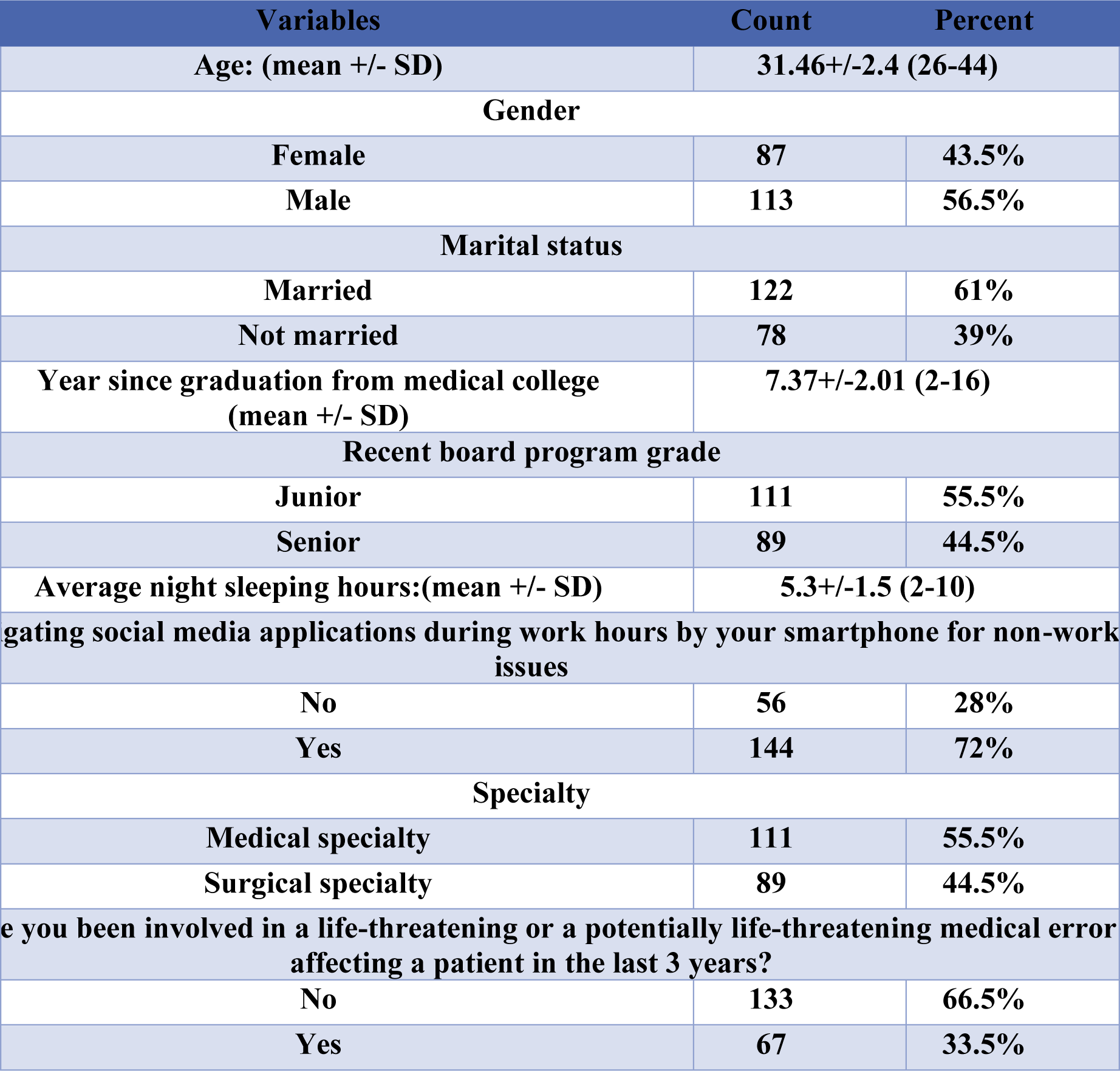
Characteristics of Participants.

One third of the participants had been involved in a life threatening error potentially affecting the life of a patient within the last 3 years.

Table (2) and Figure (1) show the Contributing factors to the medical errors:

**Figure 1:**
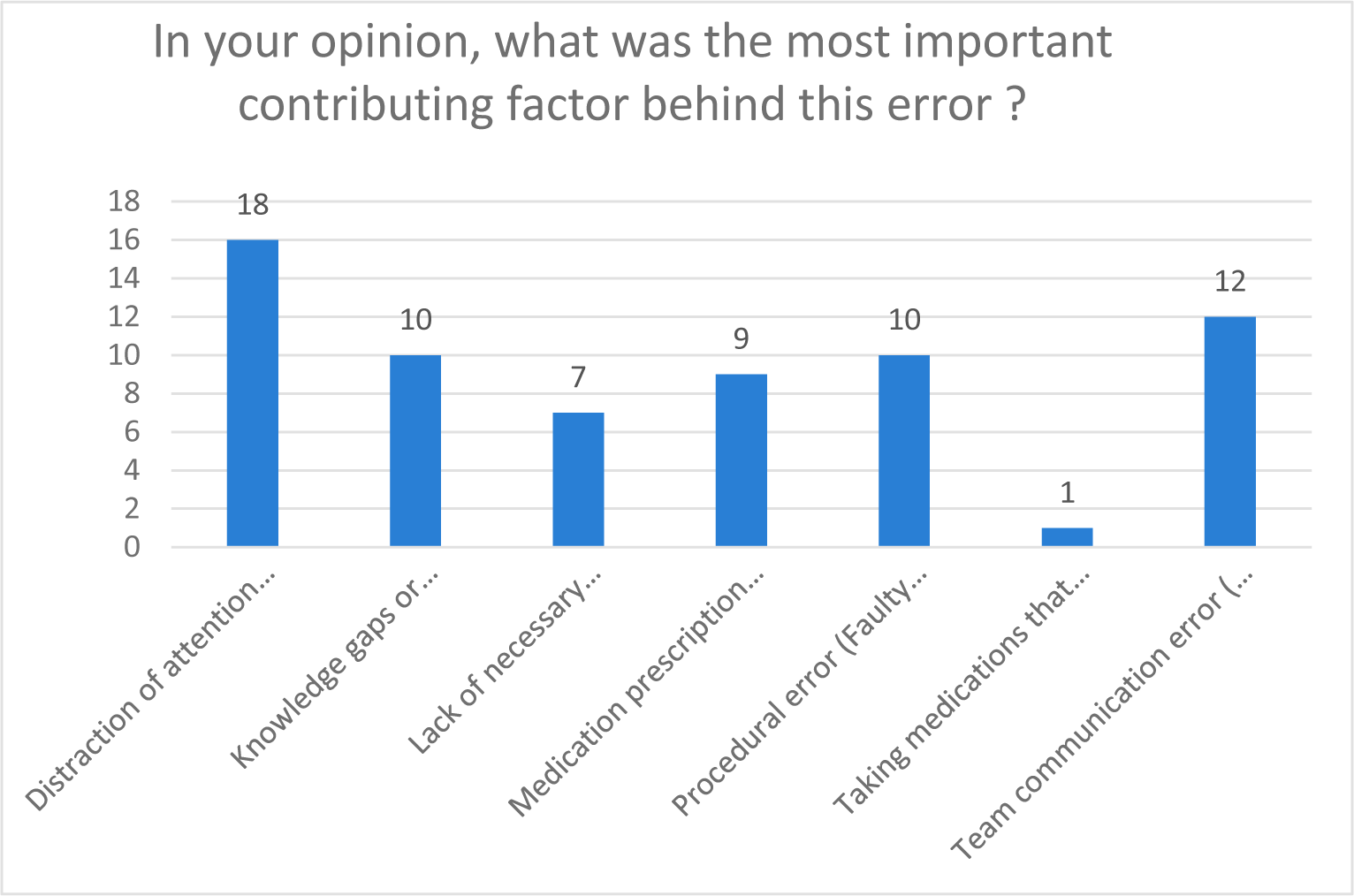
Contributing Factors Behind the Error.

**Table 2:** Choices for Contributing Factor Behind Errors.

| In your opinion, what was the most important contributing factor behind this error | Count | Percent |
| --- | --- | --- |
| Distraction of attention (From visual or auditory interference by patients' family or the teamwork members that led to the error) | 18 | 26.86 |
| Knowledge gaps or deficiency (Lack of standard guidelines, Wrong diagnosis, lack of experience etc...that contributed to the error) | 10 | 14.93 |
| Lack of necessary equipment at the time of the of the error that led to the error. | 7 | 10.45 |
| Medication prescription error (Wrong dose, Wrong abbreviation, Inappropriate hand writing) | 9 | 13.43 |
| Procedural error (Faulty technique when doing a procedure that led to the error) | 10 | 14.93 |
| Taking medications that cause sedation (tranquilizers, antihistamine, antidepressants...) | 1 | 1.49 |
| Team communication error (inadequate or unclear order from you to junior doctor or nurses which led to the error). | 12 | 17.91 |

Among participants who have made medical errors, distraction of attention (From visual or auditory interference by patients’ family or the teamwork members that led to the error) was the cause of their error accounting for 26.86% (18 individual).

Knowledge gaps or deficiency (Lack of standard guidelines, Wrong diagnosis, lack of experience etc.…that contributed to the error) accounted for 14.93% (10 individual), while lack of necessary equipment at the time of the of the error that led to the error accounting for 10.45% (7 individuals).Also 13.43% (9 individuals) chose that Medication prescription error (Wrong dose, Wrong abbreviation, Inappropriate hand writing).

Procedural error (Faulty technique when doing a procedure that led to the error) accounted for 14.93% (10 individuals).Taking medications that cause sedation (tranquilizers, antihistamine, antidepressants…) accounted for 1.49% (1 individual).

Team communication error (inadequate or unclear order from you to junior doctor or nurses which led to the error) accounted for 17.91% (12 individual)

Table (3) and figure (2) provide descriptive statistics about the strongest feelings among participants following medical error. From all 67(33.5%) participants who involved in medical errors, most 20 (29.85%) experienced emotional distress, 14 (20.9%) experienced frustration, 12 (17.91%) felt guilty, 9 (13.43%) sadness, 4 (5.97%) fear, 4 (5.97%) inadequacy and 4 (5.97%) was not their fault.

**Figure 2:**
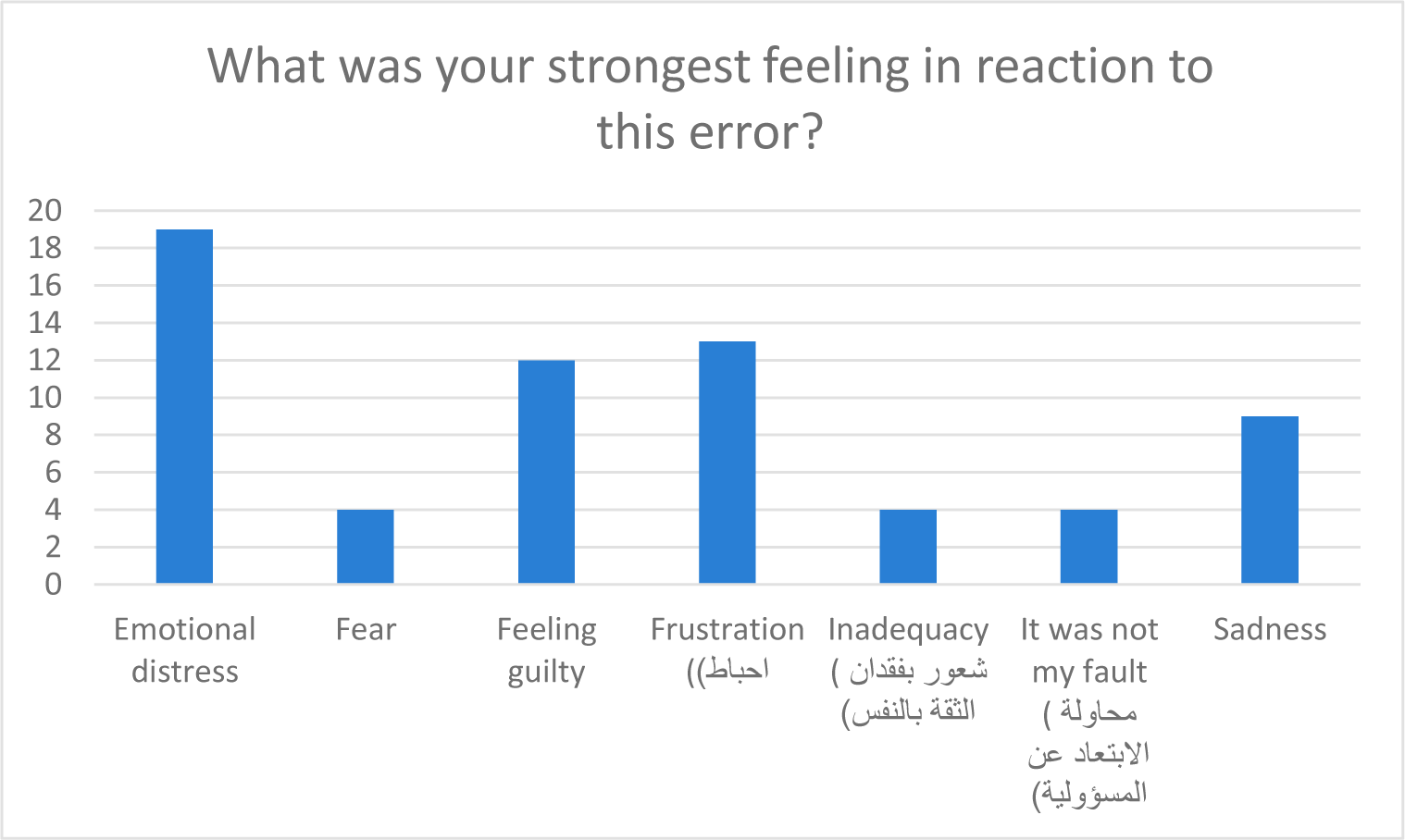
Strongest Feeling in Reaction to This Error.

**Table 3:** Choices for The Strongest Feelings.

| What was your strongest feeling in reaction to this error? | Count | Percent |
| --- | --- | --- |
| Emotional distress | 20 | 29.85 |
| Fear | 4 | 5.97 |
| Feeling guilty | 12 | 17.91 |
| Frustration | 14 | 20.90 |
| Inadequacy | 4 | 5.97 |
| It was not my fault | 4 | 5.97 |
| Sadness | 9 | 13.43 |

Table (4) and figure (3) showed the most obvious behavioral response happened after the error. From all 67 (33.5%) participants who involved in the medical errors, most 40 (58.21%) participants tend to increase vigilance (paying more attention to details of patients, decrease trust of others, always rechecking the lab report, become more careful),as well 8 (11.94%) from them increased the defensive attitude by (order more tests, keep errors to themself more often, avoid similar patients, see fewer patients). Also 19 (28.36%) from them tend to increase information seeking (always seek more advice from senior staff, ask more literature reference, read more from the book about the cases).

**Figure 3:**
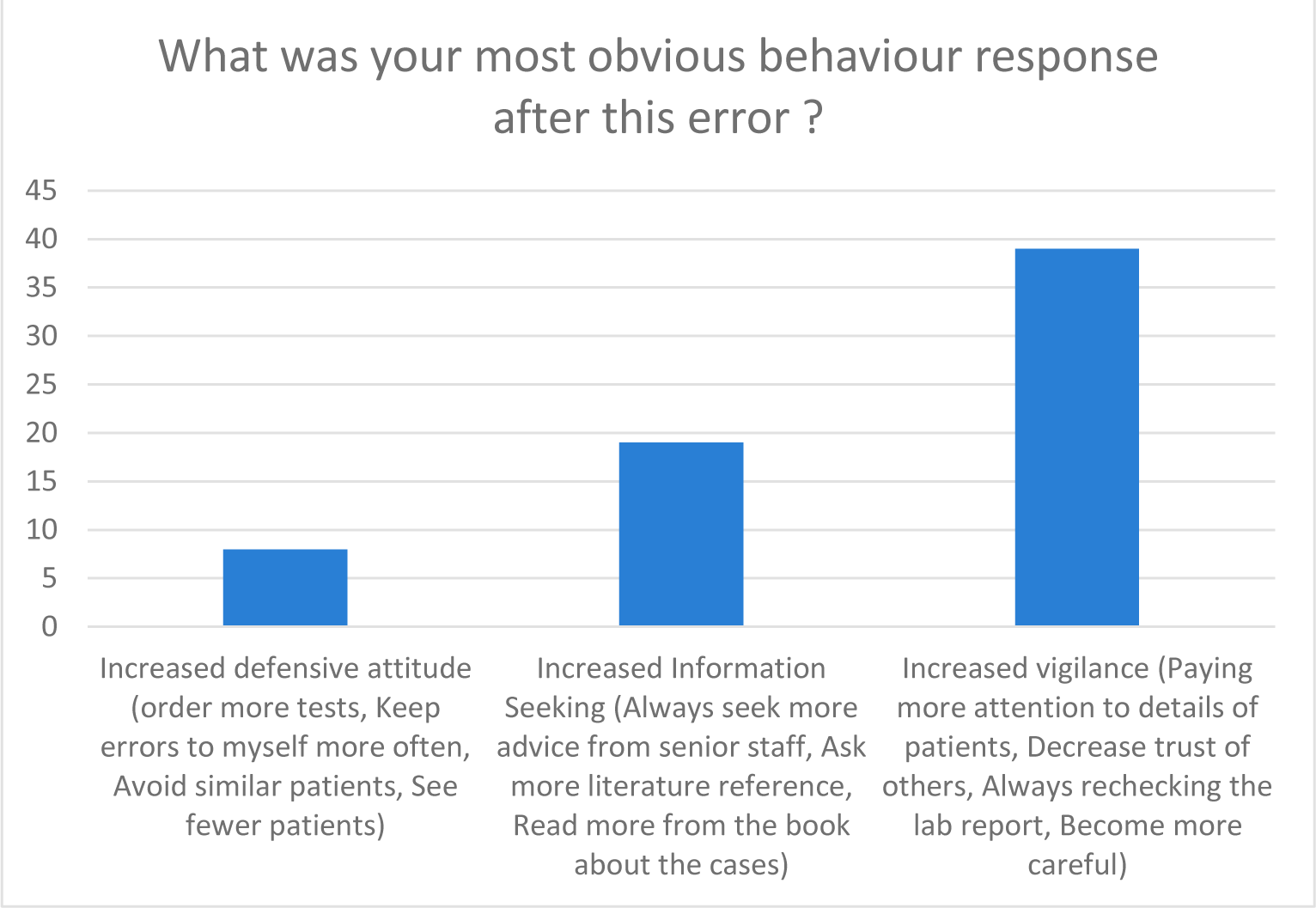
Behavioral Response Following the Error.

**Table 4:** Choices for The Behavioral Response.

| **What was your most obvious behavior response after this error?** | **Count** | **Percent** |
| --- | --- | --- |
| **Increased defensive attitude (order more tests, keep errors to myself more often, avoid similar patients, See fewer patients)** | **8** | **11.94** |
| **Increased Information Seeking (Always seek more advice from senior staff, ask more literature reference, Read more from the book about the cases)** | **19** | **28.36** |
| **Increased vigilance (paying more attention to details of patients, decrease trust of others, always rechecking the lab report, Become more careful)** | **40** | **59.70** |

Table (5) revealed a comparison between participants who were involved with those who were not involved in errors and their socio-demographic characteristics. There is statistically significant association between gender and medical errors. (P value < 0.0001). Notably, males tended to involve in medical errors more than females with a number of 46 (40.7%) compared to 21 (24.13%) respectively.

**Table 5:** Comparison of Two Groups Regarding Involvement of Errors.

| **Variables** | **Have you been involved in a life-threatening?** |  | **P value** |
| --- | --- | --- | --- |
|  | **No (n=133)** | **Yes (n=67)** |  |
| **Age (mean +/- SD) \*** | **31.4 +/- 2.3** | **31.5 +/- 2.3** | **0.77** |
| **years since graduation (mean +/- SD) \*** | **7.4 +/- 2.1** | **7.1 +/- 1.6** | **0.3** |
| average night sleeping hours (mean +/- SD) * | 5.42 +/- 1.5 | 5.2 +/- 1.4 | 0.318 |
| Gender ** |  |  |  |
| Female (n=87) (100%) | 66 (75.86%) | 21 (24.13%) | 0.0001 |
| Male (n=113) (100%) | 67 (59.29%) | 46 (40.7%) |  |
| Marital status ** |  |  |  |
| Married (n=122) | 80 (65.5%) | 42 (34.4%) | 0.12 |
| Not married (n=78) | 53 (67.94%) | 25 (32.05%) |  |
| Recent board program grade ** |  |  |  |
| Junior (n=111) (100%) | 73 (65.7%) | 38 (34.23%) | 0.06 |
| Senior (n=89) (100%) | 60 (67.4%) | 29 (32.58%) |  |
| Using social media applications during work hours by your smartphone for non-work issues ** |  |  |  |
| No (n=56) (100%) | 33 (58.9%) | 23 (41.09%) | 0.0001 |
| Yes (n=144) (100%) | 100 (69.4%) | 44 (30.5%) |  |
| Specialty ** |  |  |  |
| Medical specialty (n=111) (100%) | 78 (70.2%) | 33 (29.7%) | 0.0001 |
| Surgical specialty (n=89) (100%) | 55 (61.7%) | 34 (38.2%) |  |

*Student-t test was used.*
*\*\* Chi square test was used.*

In addition, there is statistically significant association between decreased navigating social media applications during work hours for non-work and medical error (P value < 0.0001).

Furthermore, there is statistically significant association between specialty and medical errors (P value < 0.0001) as surgical specialty tended to involve in medical errors more than medical specialty with a number of 34 (38.2%) compared to 33(29.7%) respectively.

Non-significant difference in the means of age (P value = 0.77), year since graduation from medical college (P value = 0.3), average night sleeping hours (P value = 0.318) and medical errors was found.

Married participant 42 (34.4%) tend to involve in medical errors more than non-married participants 25 (32.05%). However, there is no statistically significant association between marital status and medical errors (P value = 0.12).

Moreover, Junior doctors 38 (34.23%) tend to involve in medical errors more than senior doctors 29 (32.58%). However, there is no statistically significant association between board program grade and medical errors (P value = 0.06).

Table (6) shows a comparison within medical and surgical specialties and their socio-demographic characteristics. In this study females in medical specialty which account for 11 individuals (16%) tend to be involve in medical errors more than the females in surgical specialty 10 individuals (15%) unlike males who were more involved in the surgical specialty 24 (36%) more than the medical specialty 22(33%). However, there is no statistically significant association between gender and specialty of participants who involved in medical errors (P value = 0.729)

**Table 6:** Comparison Within the Group with Medical Errors.

| Variables |  | Specialty |  |  |  | P value |
| --- | --- | --- | --- | --- | --- | --- |
|  |  | Medical specialty (n=33) |  | Surgical specialty (n=34) |  |  |
|  |  | Count | Percent | Count | Percent |  |
| Gender** | Female (n=21) | 11 | 16% | 10 | 15% | 0.729 |
|  | Male (n=46) | 22 | 33% | 24 | 36% |  |
| Age* |  | 31.21+/-2.1 |  | 31.94+/-2.8 |  | 0.23 |
| Marital status** | Married (n=42) | 20 | 30% | 22 | 33% | 0.729 |
|  | Not married (n=25) | 13 | 19% | 12 | 18% |  |
| Average night sleeping hours* |  | 5.58+/-1.6 |  | 4.94+/-1.25 |  | 0.07 |
| Usage of social media applications during work** | No (n=23) | 14 | 21% | 9 | 13% | 0.16 |
|  | Yes (n=44) | 19 | 28% | 25 | 37% |  |
| Time of occurrence of the error** | Day time (n=22) | 9 | 13% | 13 | 19% | 0.4 |
|  | can't recall (n=14) | 6 | 9% | 8 | 12% |  |
|  | Night time (n=31) | 18 | 27% | 13 | 19% |  |

\* Student-t test was used.
\*\* Chi square test was used.

Non-significant difference in the means of age (P value = 0.23), average night sleeping hours (P value = 0.07) and specialty of participants who involved in errors was found.

Moreover, married doctors in medical specialty 20 (30%) tend to involve in medical errors less than married doctors in surgical specialty 22(33%) unlike the non-married doctors who were 13 (19%) in medical specialty and 12(18%) in surgical specialty. However, there was no statistically significant association between marital status and specialty of participants who involved in errors (P value = 0.729),

Doctors who do not navigate social media during work in medical specialty 14(21%) tend to be involved in errors more than those in surgical specialty 9(13%), in contrast doctors who navigate social media 19(28%) in medical specialty tend to involve in errors less than those 25(37%) in surgical specialty. However, there was no statistically significant association between navigation social media applications during work and specialty of participants who involved in errors (P value = 0.16),

According to the time of occurrence of errors, the majority of the participants in medical specialty tend to be involved in medical errors at night time 18(27%) more than those in surgical specialty, in contrast other participants in surgical specialty tend to be involved in errors during day time 13(19%) more than those in medical specialty 9 (13%), the rest of participants did not recall the time of occurrence of error by 6(9%) in medical specialty and 8(12%) in surgical specialty. However, there was no statistically significant association between time of occurrence of the error and the specialty of participants who involved in such error.

## DISCUSSION

In this study 33.5% (67 individual) reported their involvement in serious medical errors. Simila findings were found in three different studies, which showed major errors resulting in death in 31%, 34% and 39% respectively ^(33-35)^.

After studying **the most important contributing factors behind the medical errors** in this study, the most common cause that was chosen by the participants was distraction of attention (From visual or auditory interference by patients…) and the second common cause is team communication error (inadequate or unclear order from you to junior doctor or nurses which led to the error). A similar study found that distractions during surgical procedures are associated with team inefficiency and medical error and it also mentioned that the most distracting elements in the operation area were the equipment sounds. With the exception of music, every possible auditory distraction in this study was linked to a higher proportion of a certain degree of detrimental effect on the operation’s flow. The equipment and environment contained the top 5 distractions. The top 5 most frequent distractions were consistently phone calls, pagers, beepers, and case-related correspondence. The top four factors that were thought to have a favorable influence on the surgical process were teaching, consultation, music, and case-relevant communications. larger ratings for “bothersome” distractions seemed to be associated with a larger degree of perceived detrimental effect on the surgical process. The least distracting element was vision, which also seemed to cause minimal positive impact on the flow of surgery ^(36)^.

A similar study said that teamwork is the mainstay of health care facilities. Promoting the development of communication skills is essential for safe, compassionate, and high-quality care. Such commitment ensures the dialog and interaction between medical professionals and other disciplines, which will inevitably affect the wellbeing of the patients and their families ^(37)^. One of the main obstacles to providing safe and high-quality healthcare in a medical facility is working alone and having little or no communication inside and among teams ^(38)^. In actuality, poor team communication with patients and their families results in more than half of serious injuries and fatalities ^(39,40)^.

As a consequence of medical error, all participants reported **negative emotions followed their involvement in medical error**, most of participants experienced emotional distress, the rest experienced frustration, guilty, sadness, fear, inadequacy and some of them admit it was not their fault. Similar feelings were noted in different study ^(32)^. Although, it should consider as serious issue not only because these personal feelings of making error can be profound but also because in many studies these negative feelings can result in decreasing of patient care ^(41,42)^. Putting together it suggests a vicious cycle whereby medical errors may lead to personal distress, which then contributes to further deficits in patient care ^(43)^.

Medical errors result in **significant changes in participants’ behavioral learning**. this study showed that most participants tend to increase vigilance by paying more attention to details of patients, decrease trust of others, always rechecking the lab report, become more careful. Other study suggest that there are a number of effective strategies to increase physician vigilance after medical errors. These strategies include CIRS (Critical incident reporting systems), simulation training, RCA (Root cause analysis), and feedback and debriefing. These strategies can help to reduce the frequency and severity of medical errors and improve patient safety ^(44)^. The second most behavioral response that participants have choose in this study is increase information seeking (always seek more advice from senior staff, ask more literature reference, read more from the book about the cases). However, little research have examined the impact of medical errors on physicians’ information-seeking habits. A study showed that physicians who had experienced a medical error were more likely to engage in information-seeking behavior than those who had not experienced an error. Specifically, physicians who had experienced an error were more likely to seek information from colleagues, journals, and online resources. They were also more likely to believe that information-seeking was important for preventing medical errors ^(32)^. The third behavioral response was increase defensive attitude There is a study about how doctors may increase their defensive attitude after medical errors in a number of ways, including ordering more tests, doctors may order more tests to rule out potential problems, even if the tests are not clinically indicated, avoiding similar patients that doctors may avoid seeing patients who have similar characteristics to patients they have made mistakes with in the past and practicing defensive documentation, doctors may document their care in a way that is designed to protect them from lawsuits, even if the documentation is not accurate ^(45)^.

This study showed **gender** is significantly associated with medical error in a manner that male doctors tended to involve in medical errors more than female doctors. Different study shows male doctors in the United States were three times more likely than women to have claims for malpractice made against them ^(46)^. This could be because women have longer consultations, are more patient centered, engage in more emotionally focused talk, counsel more psychosocially, and that their patients speak more ^(47)^.

Furthermore, there is statistically significant association between **decreased navigation social media applications** during work hours for non-work issues and medical error, in spite knowing that in this study the most contributing factor behind medical errors is distraction of attention, this paradox can be explained as distractions may come from external source for example social media or internal source in form of internally generated distractions such as task unrelated thoughts or mind-wandering during task performance, thus it have been associated with impaired performance of the task ^(48)^, as a result medical errors may occur. Or unfortunately this paradox could be because of the small sample size being studied due to restriction of time, effort and team size which clearly affect research results. A third possible explanation is that doctors who are distracted by the social media navigation share poorly in caring for patients. Further research are needed.

**Specialty** is significantly associated with medical errors in manner that participants in surgical specialty tended to involve in medical errors more than medical specialty. Another study showed the same finding with percentage of 45.5% of medical errors occurred in surgical departments and 14.1% of errors occurred in internal medicine departments ^(49)^. This could be due to insufficient clinical experience of the surgeons and poor operation skills are partly responsible for medical errors^(50)^. While medical specialty involved in medical errors due to inexperience of medical staff, as well as poor communication between doctors and patients ^(51)^, Even that research found internal medicine department is major area of disputes^(52)^, probably these disputes could be over the diagnosis.

In this study, junior doctors tend to be involved in medical errors more than senior doctors. A study showed that patient care was compromised when juniors did not access senior help, often when working outside their usual team environment. Lack of cooperation between teams and poor continuity of care also contributed to errors ^(53)^. Another study show junior doctors were 1.57 times more likely to cause an error than senior staff ^(54)^. However, there is no statistically significant association between board program grade and medical errors.

Correspondingly, there is non-significant difference in the means of age and medical errors. Another study showed doctors who are in specialty training or not in specialty training but with less experience make more prescribed errors than consultants ^(55)^.

Non-significant difference was found in means of average night sleeping hours and medical errors but another study done in Saudi Arabia found that there is direct relationship between sleep deprivation and medical errors and was found to be significant ^(56)^.

In this study we found there is no statistically significant association between average night sleeping hours and specialty of participants who involved in errors, but another study has demonstrated a strong association between sleep deprivation and medical errors among participants in surgical specialties. It shows that sleep-deprived surgeons are more likely to commit technical errors during procedures, make poor decisions, and experience increased fatigue, which can compromise patient safety. Studies suggests that sleep-deprived medical professionals are more prone to diagnostic errors, medication errors, and adverse patient outcomes ^(57)^, this discrepancy between our study and other studies could be due to unfortunately the small sample size, effort and team size which clearly can affect research results.

We found that there is no statistically significant association between navigation social media applications during work and specialty of participants who involved in errors. However, similar study suggests that social media use during work hours may pose a potential risk for medical errors, particularly among medical professionals. The distractions and multitasking associated with social media use may impair focus, concentration, and adherence to established protocols, increasing the likelihood of errors. Surgical professionals, who tend to engage less in social media during work hours, may experience fewer distractions and maintain a higher level of focus during critical tasks, potentially contributing to their lower error rates ^(58)^.

Finally, in this study the majority of the participants in medical specialty tend to be involved in medical errors at night time more than those in surgical specialty, in contrast other participants in surgical specialty tend to be involved in errors during day time more than those in medical specialty. This may be due to a number of factors, such as the different types of errors that occur in each specialty, the different staffing levels that are typically available during the day and night, and the different workflow patterns that are used in each specialty ^(59)^.

## CONCLUSION

Distraction of attention was the most contributing factor behind medical errors. Emotional distress was the most common feeling experienced by participants. Increasing vigilance was the most behavioral response among participants following their involvement in medical errors. Furthermore, male doctors tend to be involved in medical errors more than female doctors. Surgical specialty tends to be involved in medical errors more than medical specialty.

## LIMITATIONS

- One of the most important limitations of our research is the small sample size and the small number of hospitals being studied, due to the restriction of time, effort, and team size, which clearly affects the research results.
- Among other things, the questionnaires were completed by self-reporting, which may lead to over- or under-declaration.

## RECOMMENDATIONS

- Attention should be paid to the medical errors with regular evaluations of their effects on patient outcomes, healthcare costs, doctors and overall quality of healthcare delivery.
- Teaching medical students how to disclose a medical error.
- Address the importance of identifying and preventing errors, highlighting initiatives, and technologies that can improve patient safety.
- Number of effective strategies to increase physician vigilance after medical errors. These strategies include CIRS (Critical incident reporting systems), simulation training, RCA (Root cause analysis), and feedback and debriefing and thus improve patient safety.

## Data Availability

All data produced in the present study are available upon reasonable request to the authors

